# Evaluation of a city-wide school-located influenza vaccination program in Oakland, California with respect to race and ethnicity: a matched cohort study

**DOI:** 10.1101/2021.07.21.21260311

**Authors:** Anna Nguyen, Benjamin F. Arnold, Chris J. Kennedy, Kunal Mishra, Nolan Pokpongkiat, Anmol Seth, Stephanie Djajadi, Kate Holbrook, Erica Pan, Pam D. Kirley, Tanya Libby, Alan E. Hubbard, Arthur Reingold, John M. Colford, Jade Benjamin-Chung

**Author notes:** **Corresponding Author:** Anna Nguyen, 259 Campus Drive 19 HRP Redwood Building T152A, Stanford University, Stanford, CA 94305.

## Abstract

**Objectives:** To evaluate the effectiveness of city-wide school-located influenza vaccination by race/ethnicity from 2014-2018.

**Methods:** We used multivariate matching to pair schools in the intervention district in Oakland, CA with schools in West Contra Costa County, CA, a comparison district. We estimated difference-in-differences (DIDs) in caregiver-reported influenza vaccination coverage and laboratory-confirmed influenza hospitalization incidence.

**Results:** Differences in influenza vaccination coverage in the intervention vs. comparison site were larger among White and Latino students than Asian/Pacific Islander (API), Black, and multiracial students. Concerns about vaccine effectiveness or safety were more common among Black and multiracial caregivers; logistical barriers to vaccination were more common among White, API, and Latinos. In both sites, hospitalization in 2017-18 was higher in Blacks vs. other races/ethnicities. All-age influenza hospitalization incidence was lower in the intervention site vs. comparison site among White/API individuals in 2016-17 and 2017-18 and Black older adults in 2017-18, but not in other groups.

**Conclusions:** SLIV was associated with higher vaccination coverage and lower influenza hospitalization, but associations varied by race/ethnicity. SLIV alone may be insufficient to ensure equitable health outcomes for influenza.

## Background/Context

To reduce influenza transmission, the Advisory Committee on Immunization Practices (ACIP) recommends annual influenza vaccinations for all Americans over 6 months of age, with a target coverage level of 80% in non-institutionalized, non-elderly persons.^1^ During recent influenza seasons, all racial/ethnic groups experienced low vaccination levels that fell short of this goal, with communities of color having the lowest coverage. While 49% of white adults received an influenza vaccine during the 2018-19 season, only 39% of Black adults, 37% Hispanic adults, and 44% of Asian adults were vaccinated.^2^

Racial/ethnic disparities in vaccination coverage can be attributed to non-belief in the utility of vaccinations, institutionalized racism, and distrust of medical institutions that contribute to vaccine hesitancy among communities of color.^3–7^ These inequities contribute to disproportionately high influenza morbidity rates among disadvantaged racial/ethnic groups, resulting in elevated rates of hospitalization and death. Prior research on the social determinants of influenza hospitalization showed that Black/African Americans and Hispanics had higher risks of hospitalization compared to Whites.^8–10^ Differences in hospitalizations by race/ethnicity are linked to other socioeconomic risk factors which disproportionately impact communities of color, such as low household income or high residential density.^8–10^Increasing vaccination coverage among marginalized groups may reduce race/ethnicity inequities in influenza morbidity and mortality.

School-located influenza vaccination (SLIV) programs aim to increase vaccination coverage levels among young children by providing free vaccination in schools. SLIV has the potential to reduce barriers to vaccination that disproportionately impact communities of color. Prior community-based interventions increased influenza vaccination coverage among communities of color by reducing logistical barriers to vaccination through door-to-door and street-based immunizations.^11^ By increasing vaccination coverage, SLIV may contribute to herd immunity and reduce influenza transmission community-wide, which may reduce racial/ethnic disparities in influenza ^12,13^. Prior studies reported that SLIV programs were associated with increased influenza vaccination coverage^14–20^ and decreased school absences^14–17,20–22^ and student illness^14–17^, but no studies have measured the differential impacts of large-scale SLIV interventions by race/ethnicity.

We previously reported results from an evaluation of a city-wide SLIV program delivered in elementary schools in Oakland, California from 2014-2018.^20^ We found that the intervention was associated with 7-11 percentage points higher vaccination coverage among school-aged children and 17 to 37 lower incidence of influenza hospitalizations per 100,000 during influenza seasons in which a moderately effective vaccine was being used. Here, we investigated whether SLIV effectiveness varied by race/ethnicity in a pre-specified subgroup analysis.

## Methods

### School-located influenza vaccination intervention

Starting in 2014, the Shoo the Flu program provided free influenza vaccinations to elementary school students city-wide in Oakland, California. The program aimed to increase vaccination coverage in children and contribute to herd protection in the surrounding community. The intervention was offered to all public elementary schools, charter schools, and preschools, as well as private schools, in the city of Oakland.

All students at participating schools were eligible to be vaccinated, regardless of their insurance status. During the 2014-2018 seasons, Shoo the Flu served between 95 and 139 schools and vaccinated between 7,502 and 10,106 students (22-28% of eligible students) each year. Additional program details are reported elsewhere.^20^

In the 2014-15 and 2015-16 seasons, the program primarily provided live attenuated influenza vaccines (LAIV), with inactivated injectable influenza vaccine (IIV) available to students with contraindications. In LAIV seasons, the vaccine had relatively low effectiveness against the predominant circulating strain.^23^ As a result, in 2016-17 and 2017-18, only the IIV was offered, consistent with Advisory Committee on Immunization Practices (ACIP) recommendations, and the vaccine was moderately effective against the predominant strain of the influenza virus.^23^

### Study design

We employed a matched cohort design to evaluate the effect of the intervention on vaccination coverage and influenza hospitalization. We focused our study on public elementary schools in Oakland Unified School District (OUSD, the intervention district). We excluded private and non-district charter schools because pre-intervention data on school characteristics was not available for them.

We selected West Contra Costa Unified School District (WCCUD) as the comparison district, as it resulted in the closest school pair matches based on pre-intervention student characteristics. Additional details on the matching procedure are described in Supplement 1.

### Outcomes and Data Sources

We evaluated the association between SLIV and (1) influenza vaccination coverage among school-aged children and (2) community-wide lab-confirmed influenza hospitalization.

#### Pre-Intervention District Comparison

To compare population characteristics between the comparison and intervention districts, we obtained pre-intervention data on socioeconomic status, school enrollment, and race/ethnicity from the three-year 2013 American Community Survey (ACS).

#### Vaccination Coverage

To measure influenza vaccination coverage in the study population, we distributed two cross-sectional surveys in 22 matched school pairs ^20^A survey administered in March 2017 measured vaccination history for the 2014-17 seasons and a survey conducted in March 2018 measured vaccination history for the 2017-18 season. Sample sizes and response rates are reported in Supplement 2.

The surveys were conducted independently from the intervention. All students in participating schools were invited to participate in the surveys, regardless of their vaccination status or participation in the SLIV program. We distributed anonymous surveys to students at schools for their caregivers to report influenza vaccination status, vaccine type, and location of vaccination. Caregivers of unvaccinated students reported the reason for non-receipt of vaccine.

Caregivers self-identified by selecting from the following student races/ethnicities: White, Black or African American, Hispanic or Latino, Asian, Native Hawaiian or Pacific Islander, or American Indian or Alaska native.

#### Laboratory-confirmed influenza hospitalization

To measure potential herd effects of SLIV, we analyzed laboratory-confirmed influenza hospitalization data among all ages in the school district catchment sites. We obtained data from the California Emerging Infections Program (CEIP) in zip codes that fell within the boundaries of the intervention or comparison districts. CEIP surveillance data tracks race-specific hospitalizations for the White, Black/African American, Hispanic, Asian/Pacific Islander, Multiracial, and American Indian/Alaska Native populations. We calculated the cumulative incidence of hospitalization each season using age-, race-, and ethnicity-specific population estimates from the 2010 Census. We restricted analyses to influenza seasons using a pre-specified, data-driven definition based on local transmission patterns, as described in Supplement 3.

### Statistical analysis

Analyses were conducted in R (version 4.0.2). The pre-analysis plan, selected datasets, and replication scripts are available through the Open Science Framework (https://osf.io/v6djf/).

#### Reasons for Vaccine Non-Receipt

The 2017-18 survey included questions about reasons for non-receipt of influenza vaccine and classified each reason into the following categories: 1) logistics, 2) non-belief, 3) barriers specific to SLIV (intervention site only) (Supplement 4). We estimated the school-level prevalence of each category and summarized characteristics in schools with prevalence above and below the median prevalence in each district. We fit bivariate log-linear Poisson models to estimate the association between an indicator of whether the school fell above or below the median prevalence for the specified reason for non-receipt and school-level characteristics.^24^

#### Influenza vaccination coverage

We used linear regression models to estimate differences in influenza vaccination coverage between the intervention and comparison districts within each racial/ethnicity group, adjusting for caregiver education level. To account for clustering within matched school pairs we calculated robust sandwich standard errors, which require no assumptions about the nature of correlation within school pairs.^25^

We pre-specified subgroup analyses by race and ethnicity. Post-hoc we excluded Native American students from the stratified analyses because the group was very small (N<20 per site per year). We excluded survey responses that did not specify a student’s race/ethnicity (2017: Intervention N = 60 (2.67%), Comparison N = 90 (2.35%); 2018: Intervention N = 63 (2.60%), Comparison N = 90 (2.20%)),

#### Laboratory-confirmed influenza hospitalization

To estimate incidence ratios, we fit log-linear modified Poisson models with an offset for population size.^24^ To account for pre-intervention differences in influenza hospitalization incidence between districts, we estimated the difference-in-differences (DID) in cumulative incidences per 100,000 individuals. We defined DIDs as the difference in pre-intervention (2011-2013) and intervention period (2014-2018) incidence differences in each site. Pre-intervention trends were similar between sites.^20^

We pre-specified stratification by age groups (non-elementary school aged individuals (<= 4 years, >13 years) and older adults (>=65 years)). Because the number of elementary school aged children who were hospitalized for influenza was small, there was inadequate statistical power to estimate associations separately for this age group.

After examining the survey data, we combined White and Asian/Pacific Islander racial categories due to low incidence among Asian/Pacific Islanders. We excluded Multiracial and American Indian/Alaska Native groups from the analysis due to rare outcomes We excluded hospitalization records that did not report race/ethnicity.

## Results

Prior to the intervention, the intervention and comparison districts had generally similar demographic characteristics (Table 1). However, relative to the comparison district, the intervention districts had a higher proportion of Black/African American residents (Intervention: 26%, Comparison: 17%), but a lower proportion of White (Intervention: 41%, Comparison: 48%), Asian (Intervention: 16%, Comparison: 19%), and Hispanic or Latino (Intervention: 26%, Comparison: 33%) residents. The intervention district had a lower median household income than the comparison district, but a higher proportion of residents holding a bachelor’s degree or above.

**Table 1:** Pre-intervention characteristics of the intervention (Oakland, California) and comparison (West Contra Costa County, CA) district catchment areas for the three-year period between 2011-2013. Table reproduced from Benjamin-Chung et al (2020).^20^ Data source: 2013 American Community Survey for the 3-year period between 2011-2013

| Characteristic | Intervention (95% CI) | Comparison (95% CI) |
| --- | --- | --- |
| Median household income (dollars) | 51849 (50460, 53238) | 61596 (59662, 63530) |
| Households below the poverty level (%) | 21 (20, 22) | 15 (13, 16) |
| Highest education level (%) |  |  |
| Less than high school | 16 (15, 18) | 14 (12, 17) |
| High school graduate | 24 (21, 26) | 30 (25, 34) |
| Some college or Associate's | 46 (43, 48) | 50 (46, 55) |
| Bachelor's degree or higher | 15 (12, 17) | 6 (4, 8) |
| Children attending kindergarten in private versus public schools (%) |  |  |
| Public kindergarten | 87 (81, 92) | 86 (80, 92) |
| Private kindergarten | 13 (8, 19) | 14 (8, 20) |
| Children attending grade 1-4 in private versus public schools (%) |  |  |
| Public grades 1-4 | 89 (86, 92) | 84 (79, 88) |
| Private grades 1-4 | 11 (8, 14) | 16 (12, 21) |
| Children attending grade 5-8 in private versus public schools (%) |  |  |
| Public grades 5-8 | 89 (86, 91) | 87 (83, 91) |
| Private grades 5-8 | 11 (9, 14) | 13 (9, 17) |
| Race (%) |  |  |
| White | 41 (40, 42) | 48 (47, 50) |
| Black or African American | 26 (25, 27) | 17 (16, 18) |
| Asian | 16 (16, 17) | 19 (18, 20) |
| Other race | 9 (8, 10) | 8 (7, 9) |
| Native Hawaiian and Other Pacific Islander | 1 (0, 1) | 0 (0, 1) |
| Two or more races | 6 (6, 7) | 6 (5, 7) |
| Hispanic or Latino ethnicity | 26 (25, 27) | 33 (32, 35) |

### Vaccination Coverage

The race/ethnicity distributions of respondents were similar to the overall distributions in the sampled schools (Figure S5), with the exception of Black/African American students, who were underrepresented in both districts (Comparison: 19% in target population versus 10% in survey; Intervention: 33% in target population versus 16% in survey). The distributions of student race/ethnicity as reported in caregiver surveys are described in Table S6. In both districts, most multi-racial students identified as part Black/African American (2017: Comparison 26%, Intervention 39%) or Asian/Pacific Islander (2017: Comparison 35%, Intervention 37%) (Table S7).

Vaccination coverage levels varied by race/ethnicity, when controlled for highest caregiver education (Figure S8). In all seasons, we observed lower vaccine coverage among Black/African American and multiple race students across both districts. During the 2017-18 season, 40% (95% CI 24%, 59%) of Black/African American students were vaccinated, compared to 65% (95% CI 57%, 72%) of Asian/Pacific Islander and 46% (95% CI 28%, 66%) of White students in the comparison district. In the intervention district, 46% (95% CI 30%, 64%) of Black/African American students were vaccinated, compared to 74% (95% CI 65%, 80%) of Asian/Pacific Islander and 67% (95% CI 48%, 82%) of White students.

Associations between SLIV and vaccine coverage varied between racial/ethnic groups (Figure 1, Table S9). In the first two seasons of SLIV there were no differences in vaccine coverage for any group, apart from Asian/Pacific Islanders. In the 2016-17 season, there were increases in vaccine coverage in all groups other than multiple race students. We observed higher coverage levels in the intervention district relative to the comparison district among Black/African American (9% higher in intervention versus comparison; 95% CI 2%, 17%), Latino students (11%; 95% CI 5%, 17%),White (7%; 95% CI -1%, 14%), and Asian/Pacific Islander (5%; 95% CI 0%, 9%) students.

**Figure 1:**
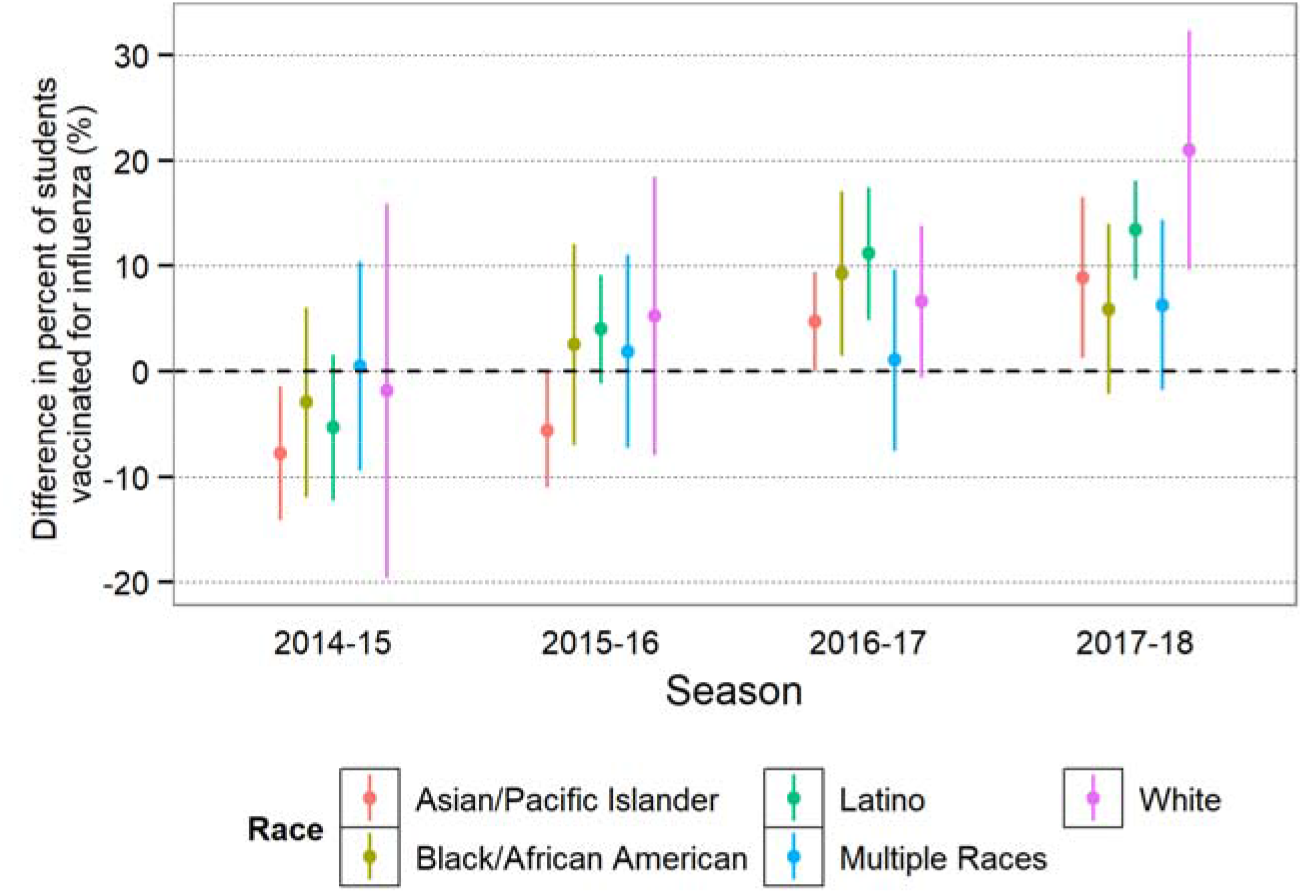
Differences in vaccination coverage among students enrolled in the intervention versus comparison district between 2014-2018. Difference in the percent of students vaccinated in the intervention district and percent of students vaccinated in the comparison district, adjusted for highest parental education level. Estimates calculated from caregiver surveys in March 2017 (for the 2014-15, 2015-16, 2017-18 seasons) and March 2018 (for the 2017-18 season). Standard errors calculated with respect to school-level clustering.

In the 2017-18 season, we observed significantly higher influenza vaccine coverage levels among Asian/Pacific Islander (9%; 95% CI 1%, 16%), Latino (13%; 95% CI 9%, 18%), and White (21%; 95% CI 10%, 32%) students in the intervention district relative to the comparison district. Vaccine coverage was higher among Black/African American (6%; 95% CI -2%, 14%) and multiple race (6%; 95% CI -2%, 14%) students, with more vaccinated students in the intervention district.

Among caregivers of students who were not vaccinated for influenza, those of Black/African American students and multiple race students were more likely to cite non-belief as a reason for vaccine non-receipt.(Figure 2) Frequency of non-belief was slightly lower in the intervention district (Black/African American: 68.5%, Multiple: 72.9%) than in the comparison district (Black/African American: 85.1%, Multiple: 76.6%). Caregivers of White, Latino, and Asian/Pacific Islander students in the comparison district more commonly reported logistical barriers to vaccination (White: 44.60%, Latino: 38.6%, Asian/Pacific Islander: 39.5%). These barriers were reported less often in the intervention district in these groups (White: 25.6%, Latino: 24.6%, Asian/Pacific Islander: 30.1%). (Figure 2). SLIV-specific concerns were most common among Asian/Pacific Islander (29.5%) and Latino (28.2%) students in the intervention district.

**Figure 2:**
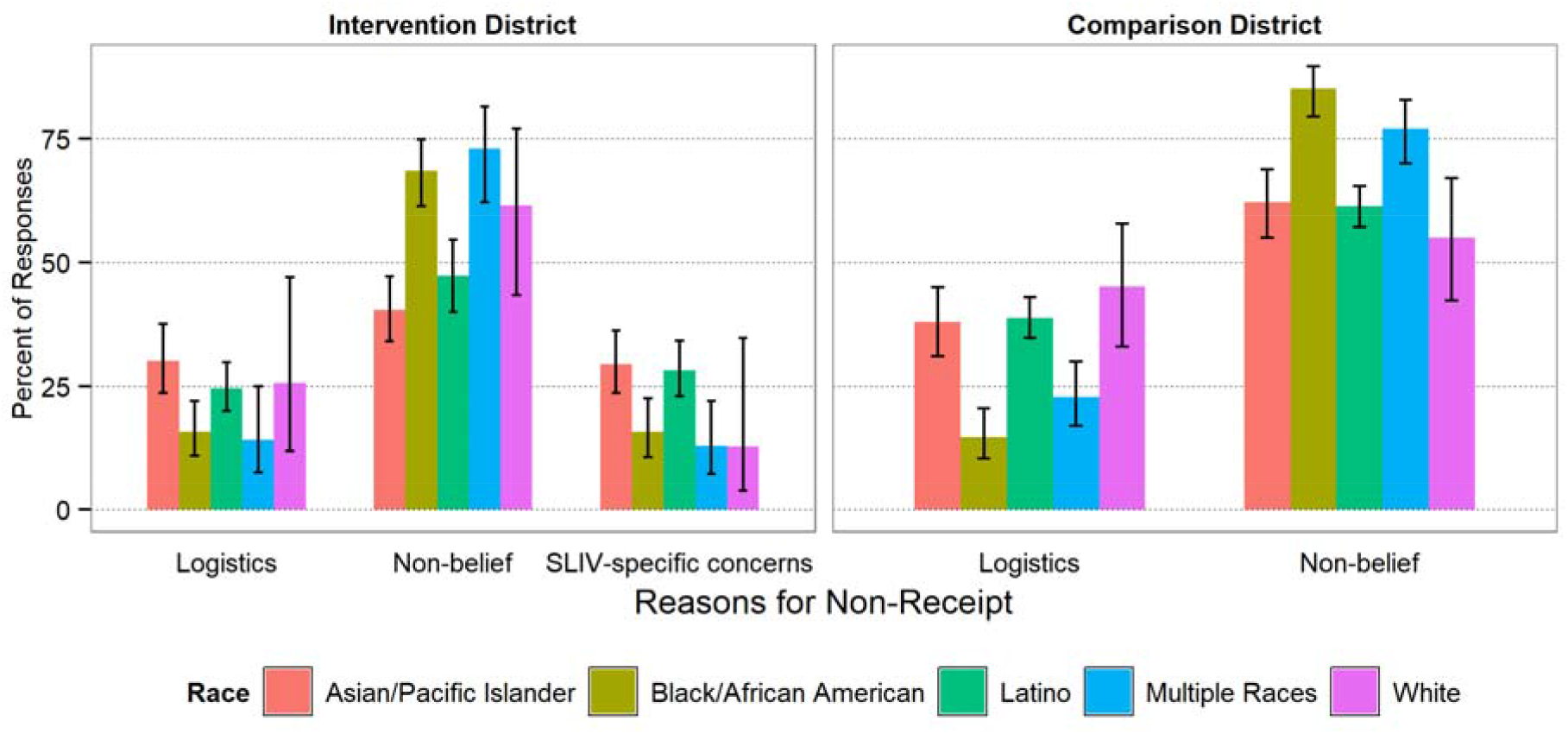
Caregiver-reported reasons for non-receipt of influenza vaccination among students during the 2017-18 season. Estimated percent of caregivers of non-vaccinated children that responded with the specified reason for non-receipt within each racial/ethnic group, calculated from survey data filled out by student caregivers in March 2018 (for the 2017-18 season). Questions corresponding to each category listed in Supplement 9.

Schools with a prevalence of non-belief in influenza vaccination above the district median had a higher percentage of White, Black/African American, andmultiple race students and a lower percentage of English learners and students eligible for free lunch in both districts (Table S10). Schools with a prevalence of logistical barriers above the district median had a higher percentage of English learners and students eligible for free lunch in both districts, and a higher percentage of Hispanic students in the comparison district.

### Laboratory-confirmed influenza hospitalization

The incidence of influenza hospitalizations varied by race/ethnicity (Figure S11). These differences were most pronounced during the 2016-17 and 2017-18 seasons, which had more higher rates of influenza. In both seasons, the incidence of all-age, older adult, and non-elementary hospitalization for influenza in each district was highest among Black/African Americans. In 2017-18, the cumulative incidence of hospitalizations among Black/African Americans in the intervention district (111 per 100,000) was about three times higher than the cumulative incidence among White and Asian/Pacific Islanders (36 per 100,000). Similarly, the cumulative incidence among Black/African Americans in the comparison district (134 per 100,000) was nearly twice as high as the cumulative incidence among Whites and Asian/Pacific Islanders (73 per 100,000). (Figure S12) The cumulative incidence of influenza hospitalizations of Hispanic/Latinos was relatively low, at 32 hospitalizations per 100,000 in the intervention district and 43 per 100,000 in the comparison district.

Associations between SLIV and influenza hospitalizations varied by race/ethnicity (Figure 3, Figure S12). We observed fewer all-age influenza hospitalizations in the intervention district among Whites and Asian/Pacific Islanders in the 2016-17 season (DID of -25.1 per 100,000 between intervention and comparison districts; 95% CI -40.1, -10.1) and in the 2017-18 (−36.6; 95% CI -52.7, -20.5) season, the seasons when the influenza vaccine was moderately effective. Among older adults, we found large, protective DIDs among Whites and Asian/Pacific Islanders in the 2016-17 (−116.1; 95% CI -205.7 -26.5) and 2017-18 (−133.9; 95% CI -225.5, - 42.2) seasons and among Black/African Americans during the 2017-18 (−282.3; 95% CI -508.4, -56.1) season. Hispanic/Latinos experienced a smaller DID in all-age (hospitalizations (2017-18: -12.1; 95% CI -31.5, 7.4), but the incidence of influenza hospitalizations was lower in the intervention district relative to the comparison district.

**Figure 3:**
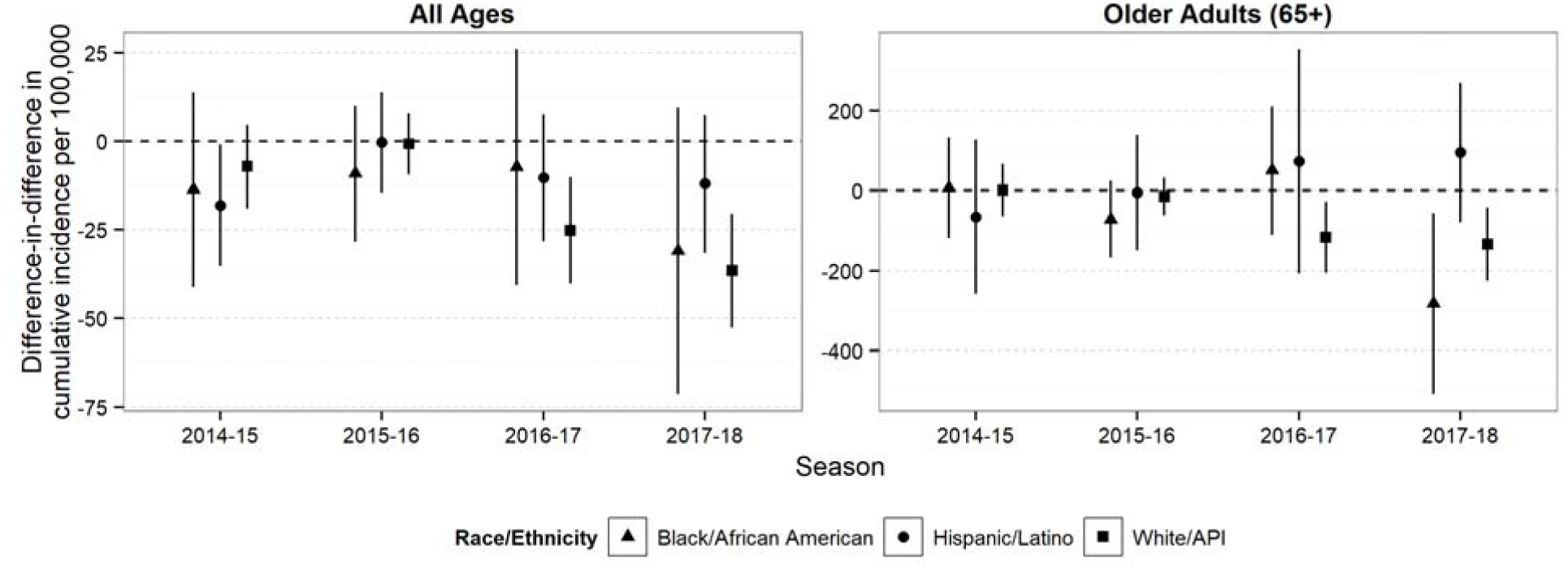
Difference-in-differences in the cumulative incidence of influenza-related hospitalizations per 100,000 in the intervention district versus comparison district, by age group and race. Difference in the difference in cumulative incidence per 100,000 of hospitalization between the intervention season and pre-intervention period in the intervention district versus the difference in cumulative incidence of hospitalization between the intervention season and pre-intervention period in the comparison district. Estimates calculated from influenza surveillance data provided by the California Emerging Infections Program.

## Discussion

We assessed the variation in the impact of a city-wide SLIV program on influenza vaccine coverage and influenza-related hospitalizations by race and ethnicity. By the third and fourth years of the program, SLIV was associated with higher vaccine coverage among elementary school children in all racial/ethnic groups. However, influenza vaccine coverage remained lower among Black/African American students compared to other groups. Associations between SLIV and higher vaccination coverage were largest among Latino and White students. Among unvaccinated students, fewer caregivers reported logistical barriers in the intervention district than in the comparison district across all racial/ethnic groups. Nonbelief in the influenza vaccine was over twice as common as reported logistical barriers in all groups and was most common among Black/African American and White caregivers. SLIV was associated with fewer influenza hospitalizations among Whites and Asian/Pacific Islanders in years when the influenza vaccine was moderately effective. Despite associations between SLIV and higher vaccine coverage in Black/African American over two seasons, we observed protective associations with influenza hospitalizations only in this group in older adults in one season.

SLIV has the potential to increase equity of vaccine coverage by reducing logistical barriers to vaccination that are more common in communities of color.^3,26^ Prior studies attributed low influenza vaccine coverage among Hispanics/Latinos to limited accessibility and affordability.^3,27,28^ In our study, logistical barriers were reported more common by caregivers of children in comparison district schools, where more students were English Learners or qualified for free lunch (a proxy for lower socioeconomic status). This was not the case in the intervention district and may suggest that SLIV helped reduce logistical barriers to influenza vaccination for students in schools with lower socioeconomic status. However, 22% of caregivers of unvaccinated students in the intervention district still reported logistical barriers. Taken together, these findings suggest that intervention reduced, but did not eliminate logistical barriers to vaccination.

In both districts, nonbelief in influenza vaccination was a more common reason students were not vaccinated than logistical barriers; however, non-belief was slightly less common in the intervention district. Nonbelief was common across all racial/ethnic groups but was most common among Black/African American, multiracial, and White caregivers. Non-belief stems from beliefs and historical contexts that vary by race/ethnicity. Among Black/African Americans, current institutional racism, as well as discrimination, exploitation, and abuse in medical practice and research contributes to distrust of medical institutions and vaccine hesitancy. ^6,29–32^ Vaccine hesitant White populations tend to be affluent, have low perceived severity of influenza, and experience lower incidence of severe influenza morbidity^33,34^

Our findings highlight a challenge to SLIV programs: increasing access to influenza vaccinations though school-based programs by itself does not address vaccine hesitancy. To achieve equitable improvements in influenza vaccine coverage, SLIV programs may need to be tailored to address concerns and beliefs of specific school communities. In prior studies, targeted, culturally-sensitive outreach campaigns that addressed the specific concerns and needs of communities and that empowered them to make informed health decisions on their own terms were more successful in increasing vaccination rates across groups.^11,35–40^

Prior studies have found that groups with higher influenza vaccine coverage have fewer influenza hospitalizations.^20,41^ However, in our study, even though SLIV was associated with higher vaccination coverage in Black/African Americans, the incidence of influenza related hospitalizations remained higher than that of other groups. Similarly, for Hispanics/Latinos, improvements in vaccination coverage did not translate to lower influenza hospitalization. Increases in vaccine coverage among this group in the SLIV site were smaller relative to increases for Whites and may not have been large enough to translate to lower rates of influenza hospitalization in these groups. Structural racism contributes to persistent barriers to vaccination and access to medical care in Black and Latino communities,^42–44^ and may be associated higher prevalence of comorbid conditions in these groups, which may have resulted in the disparities in the impact of SLIV on influenza hospitalization.^45,46^

### Limitations

Our study is subject to several limitations. Student influenza vaccination history was reported by caretakers and may be subject to poor recall. Measurements during the 2014-15 and 2015-16 influenza seasons had longer recall periods and may be more prone to error. There may have been differential accuracy of recall between the intervention and comparison districts because the presence of SLIV could have improved caregivers’ ability to correctly remember their child’s vaccination history. ^47,48^ Additionally, we saw higher levels of non-response among Black/African American students. If those who did not respond to the survey were also less likely to have been vaccinated, our results may have under-estimated the true disparities in vaccination between Black/African American students and other racial/ethnic groups. However, we previously standardized vaccine coverage models by school-district race distributions and did not find evidence of selection bias. ^20^ In addition, our estimates of overall vaccine coverage were consistent with national and state data reported by the United States Centers for Disease Control and Prevention. ^47,48^ Finally, changes in health care utilization over time could have influenced estimates for the incidence of influenza hospitalization.^49^ In this study, the first year of the intervention coincided with expanded coverage of preventative care under the Affordable Care Act. The matched cohort design and DID analysis controlled for time-invariant confounding variables, and it is unlikely that policy changes impacted the two study districts differently.

### Public Health Implications

A city-wide SLIV program was associated with higher influenza vaccination coverage in school age children, but racial/ethnic disparities in vaccine coverage were present. We did not observe associations between SLIV and reductions in the incidence of all-age hospitalization despite higher vaccine coverage among the Black/African American and Latino communities, although reductions in the incidence of hospitalizations among Black/African American older adults were observed in the 2017-18 season. SLIV shows promise for the delivery of other vaccines, such as SARS-CoV-2 vaccines. Black, Hispanic, and Native Americans and Alaskan Natives were more likely to experience severe COVID-19 illness than Whites and faced structural barriers to vaccination during early COVID-19 vaccination efforts.^50–52^ Vaccinating children will be crucial in protecting communities from COVID-19 outbreaks. Once COVID-19 vaccines receive approval for use in children, SLIV may be effective at increasing vaccine coverage levels and increasing equity of vaccination coverage.

## Conclusion

After four years of implementation, SLIV was associated with higher influenza vaccine coverage students during seasons with a moderately effective vaccine, with the largest differences observed among White and Latino students. Associations between SLIV and lower rates of influenza hospitalizations varied by race and ethnicity but were not consistent with the relative improvements in vaccination coverage by race and ethnicity. Overall, Black/African American students had the lowest overall influenza vaccination levels and Black/African and Hispanic communities had higher influenza hospitalization rates. Future SLIV programs may benefit from tailored campaigns to address race/ethnicity-specific beliefs, concerns, and structural factors that contribute to lower vaccine coverage.

## Data Availability

Data related to the analysis of vaccination coverage is available at on the associated OSF page. Data related to the analysis of hospitalization surveillance is not publicly available.

https://osf.io/v6djf/

https://github.com/anna-nguyen/stf-race-ethnicity

## Notes

### Competing Interest Statement

EP reports prior grant support from the Shoo the Flu organization during her work at the Alameda County Public Health Department; PDK and TL report grant support from the U.S. Centers for Disease Control and Prevention to the California Emerging Infections Program.

### Funding Statement

This evaluation was supported by the Flu Lab (https://theflulab.org/) through a grant (Award number: 20142281; PI: AR) awarded to the University of California, Berkeley. Decisions regarding study design, data collection, statistical analysis, manuscript preparation, and publication were made independently from funders.

### Author Declarations

This study was reviewed and approved by the Committee for the Protection of Human Subjects at the University of California, Berkeley (Protocols # 2014-01-5960 and 2016-12-9406).

