## Supplemental Materials for "Evaluation of a city-wide school-located influenza vaccination program in Oakland, California with respect to race and ethnicity: a matched cohort study"

### Supplement 1: Comparison District Matching Procedure

The use of a matched cohort study minimizes the influence of confounding variables on measures of association between SLIV and vaccination coverage. By matching schools in the intervention district to similar schools in the comparison district, we aim to achieve similar distributions of unmeasured and measured confounding variables between the two groups.

To select an appropriate comparison district, we compiled a list of candidate districts of all elementary schools in the San Francisco Bay Area that had 4+ elementary schools. A genetic multivariate matching algorithm was used to generate school-pairs between the intervention district and each candidate comparison district.^53^ The matching algorithm considered pre-intervention levels of mean enrollment, class size, parental education, academic performance index scores, California standardized test scores, and school-level percentage of English language learners and students eligible for free lunch.

We calculated the generalized Mahalanobis distance in each set of school-pairs and selected the comparison district with the lower average distances. The absolute values of the standardized differences were used to assess the quality of school pair matches.

The number of elementary schools in the comparison district (N=34) was large enough to ensure adequate statistical power in our vaccination coverage analysis; sample size calculations are reported elsewhere.^20^

To assess the impact of SLIV on community influenza hospitalizations, we included all hospitalizations of residents living in ZIP codes that were at least partially contained in either school district’s boundaries.

### Supplement 2: Sample Size

OUSD is a large, diverse school district. At the start of the intervention period, there were 50 district-run elementary schools serving 19,987 students and 6 district authorized charter schools serving 4,192 students. This study was conducted in public elementary schools because pre-intervention data for preschools, private schools, and charter schools was not available.

Surveys were distributed to 22 matched school pairs.

In the 2017 survey 2,246 of the 8,121 distributed surveys were returned in the intervention district and 3,824 of the 10,056 distributed surveys were returned in the comparison district. In the 2018 survey 2,421 of the 10,110 distributed surveys were returned in the intervention district and 4,086 of the 11,820 distributed surveys were returned in the comparison district.

### Supplement 3: Influenza Season Definition

We used a data-driven approach to define influenza seasons based on local transmission. We set a 2.5% threshold for the percentage of medical visits for influenza-like illness in a week, as reported by the California Department of Public Health (CDPH), to mark the start and end of an influenza season. Each year, the season started when there were two consecutive weeks that exceeded the threshold and ended when there were two consecutive weeks that dropped below the threshold.

### Supplement 4: Categorization of caregiver-reported reasons for non-receipt

Caregivers reported reasons for their child vaccine non-receipt in a survey distributed during the 2017-18 influenza season. We categorized these reasons into three overarching groups, as described below:

*Logistics*

- “It costs too much.”
- “I didn’t have time to take my student to the doctor.”
- “I thought my student needed health insurance to get it.”
- “I didn’t know where to get it.”
- “My student is afraid of needles.”

*Non-belief*

- “I don’t believe in it.”
- “I believe it might make my student sick.”
- “Our doctor did not recommend it.”

*SLIV-specific concerns*

- “I didn’t receive the consent form to get the vaccine at school.” (Intervention only)
- “I forgot to return the consent form to get the vaccine at school” (Intervention only)
- “I didn’t trust schools to vaccinate my student.” (Intervention only)
- “I didn’t want to share my insurance information on the consent form to get the vaccine at school.” (Intervention only)

### Figure S5: Distribution of student race/ethnicity among school district, sampled schools, and survey respondents.

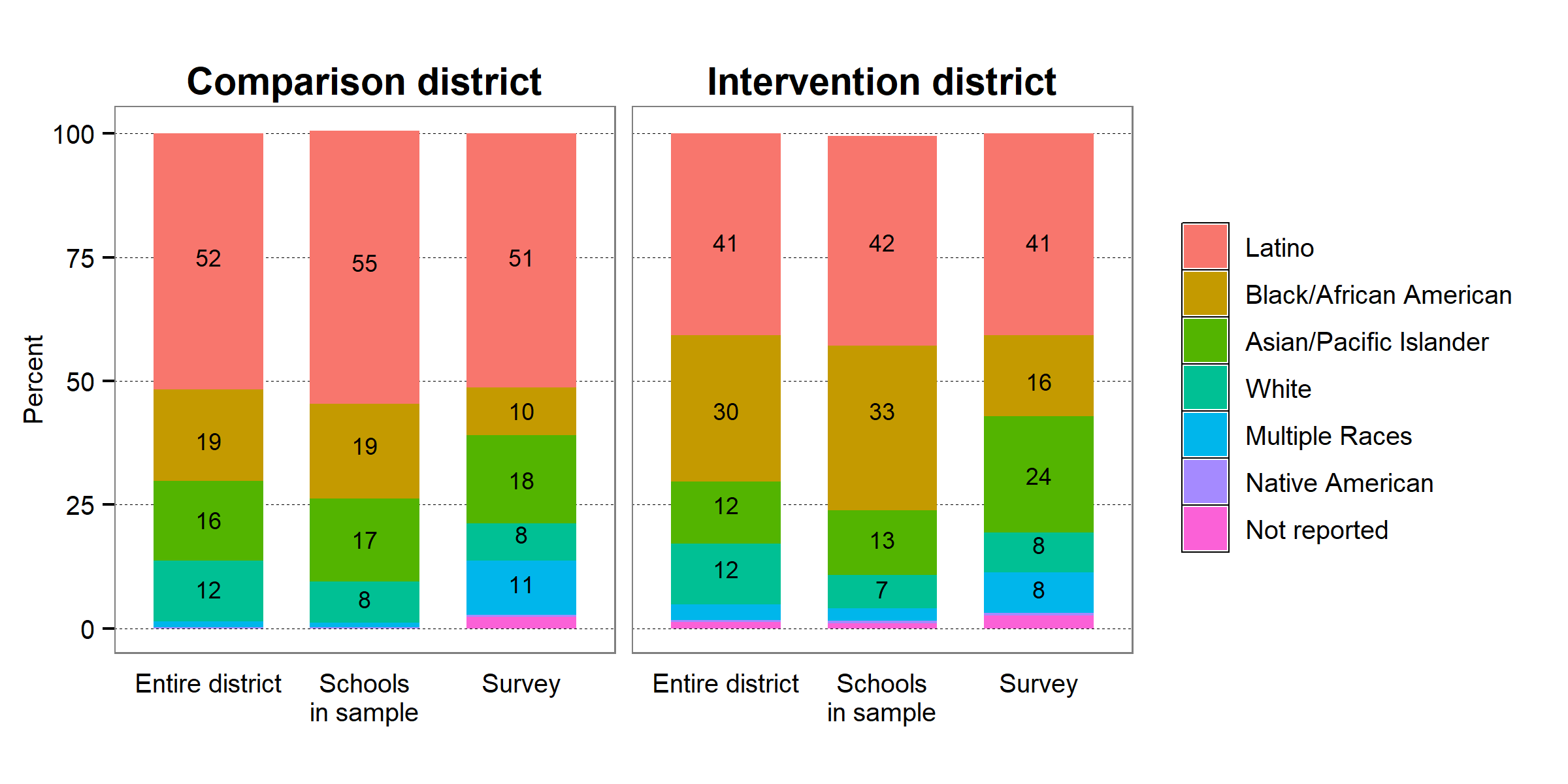

### Table S6: Distribution of student race/ethnicity reported on caregiver surveys

|  | **2017** | | | | **2018** | | | |
| --- | --- | --- | --- | --- | --- | --- | --- | --- |
|  | **Intervention** | | **Comparison** | | **Intervention** | | **Comparison** | |
| **Race/Ethnicity** | **N** | **Percent** | **N** | **Percent** | **N** | **Percent** | **N** | **Percent** |
| White | 180 | 8.0 | 292 | 7.6 | 188 | 7.8 | 324 | 7.9 |
| Black/African American | 367 | 16.3 | 371 | 9.7 | 398 | 16.4 | 402 | 9.8 |
| Latino | 916 | 40.8 | 1961 | 51.3 | 869 | 35.9 | 2055 | 50.3 |
| Asian/Pacific Islander | 529 | 23.6 | 678 | 17.7 | 643 | 26.6 | 737 | 18.0 |
| Native American | 11 | 0.5 | 17 | 0.4 | 17 | 0.7 | 24 | 0.6 |
| Multiple Races | 183 | 8.2 | 415 | 10.9 | 243 | 10.0 | 454 | 11.1 |
| Missing | 60 | 2.7 | 90 | 2.4 | 63 | 2.6 | 90 | 2.2 |

### Table S7: Distribution of race/ethnicity among multiple race students reported on caregiver surveys

|  | **2017** | | | | **2018** | | | |
| --- | --- | --- | --- | --- | --- | --- | --- | --- |
|  | **Intervention** | | **Comparison** | | **Intervention** | | **Comparison** | |
| **Race/Ethnicity** | **N** | **Percent** | **N** | **Percent** | **N** | **Percent** | **N** | **Percent** |
| Black/African American + White | 24 | 13.1 | 33 | 8.0 | 30 | 12.4 | 48 | 10.6 |
| Black/African American + Latino | 26 | 14.2 | 39 | 9.4 | 35 | 14.4 | 56 | 12.3 |
| Black/African American + API | 22 | 12.0 | 35 | 8.4 | 32 | 13.2 | 48 | 10.6 |
| Latino + White | 18 | 9.8 | 96 | 23.1 | 24 | 9.9 | 91 | 20.0 |
| Latino + API | 16 | 8.7 | 39 | 9.4 | 15 | 6.2 | 35 | 7.7 |
| API + White | 30 | 16.4 | 72 | 17.4 | 42 | 17.3 | 67 | 14.8 |
| Other | 47 | 25.7 | 101 | 24.3 | 65 | 26.8 | 109 | 24.0 |

### Figure S8: Vaccination coverage levels by race among students in the comparison and intervention districts

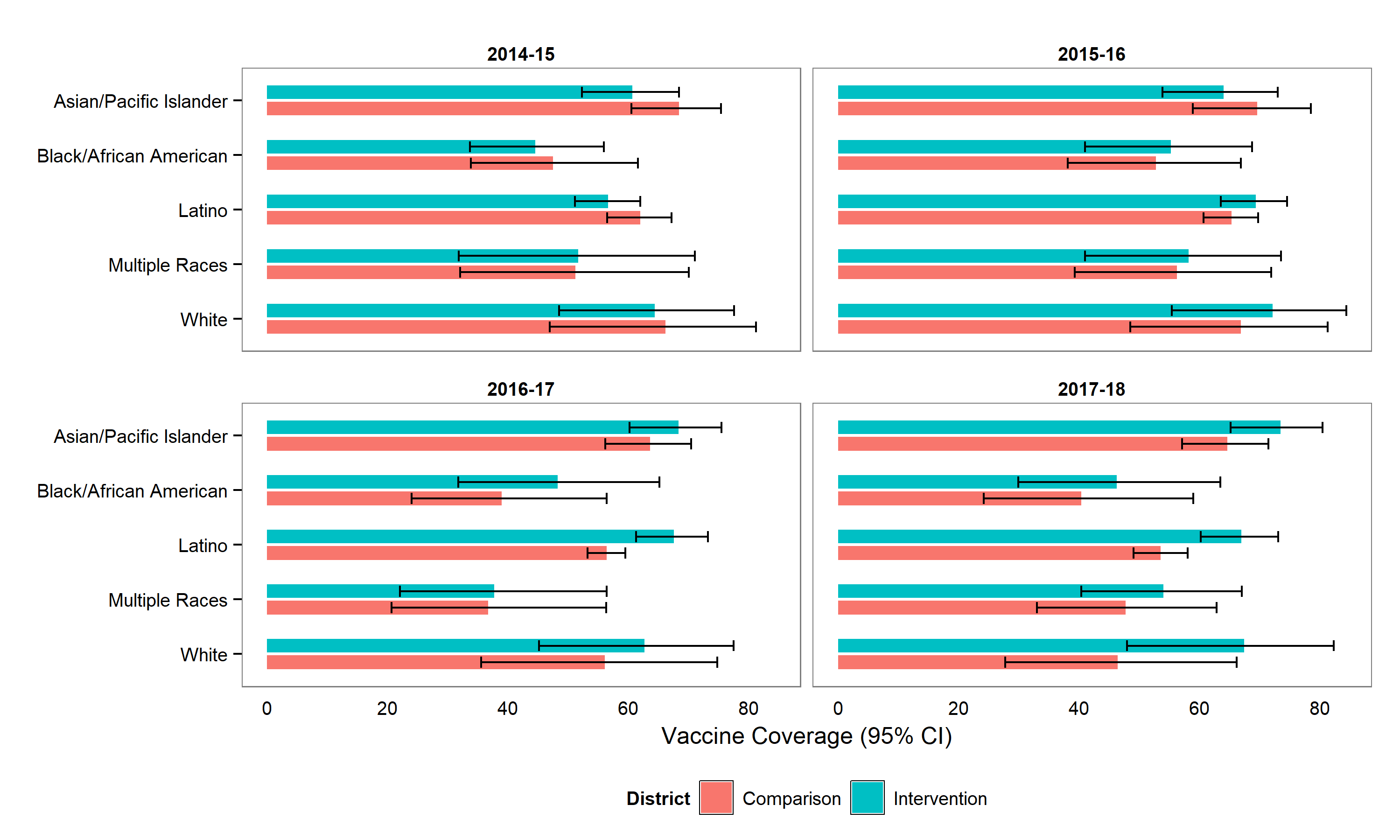

### Table S9: Differences in vaccination coverage among students enrolled in the intervention versus comparison district, adjusted for parental education.

| **Race** | **2014-15** | **2015-16** | **2016-17** | **2017-18** |
| --- | --- | --- | --- | --- |
| Asian/Pacific Islander | -7.79 (-14.12, -1.47) | -5.60 (-11.04, -0.17) | 4.74 (0.04, 9.43) | 8.87 (1.25, 16.48) |
| Black/African American | -2.94 (-11.91, 6.03) | 2.52 ( -6.97, 12.02) | 9.27 (1.51, 17.03) | 5.90 (-2.16, 13.97) |
| Latino | -5.34 (-12.22, 1.54) | 4.01 ( -1.11, 9.12) | 11.17 (4.90, 17.44) | 13.40 (8.76, 18.03) |
| Multiple Races | 0.50 ( -9.41, 10.40) | 1.89 ( -7.27, 11.06) | 1.06 ( -7.57, 9.68) | 6.27 ( -1.79, 14.34) |
| White | -1.83 (-19.57, 15.92) | 5.28 ( -7.92, 18.47) | 6.62 ( -0.59, 13.83) | 20.98 (9.65, 32.32) |

### Table S10: Distribution of school-characteristics for schools with low versus high frequencies for categories of reason for vaccine non-receipt.

|  |  | **Non-Belief** | | | **Logistics** | | |
| --- | --- | --- | --- | --- | --- | --- | --- |
|  | Characteristic (%) | Below Median | Above Median | P-Value | Below Median | Above Median | P-Value |
| **Intervention District** | White | 2.39 | 10.86 | <0.01 | 10.21 | 3.04 | <0.01 |
|  | API | 16.57 | 11.07 | <0.01 | 8.15 | 19.49 | <0.01 |
|  | Hispanic | 48.66 | 36.1 | <0.01 | 40.81 | 43.96 | 0.26 |
|  | Black/African American | 30.04 | 36.57 | 0.01 | 36.46 | 30.15 | 0.01 |
|  | Multiple race | 1.31 | 3.51 | <0.01 | 2.89 | 1.93 | 0.15 |
|  | Eligible for Free Lunch | 87.18 | 62.82 | <0.01 | 67.68 | 82.32 | <0.01 |
|  | English Learner | 48.49 | 30.73 | <0.01 | 34.25 | 44.97 | <0.01 |
| **Comparison District** | White | 7.58 | 8.77 | 0.34 | 9.41 | 6.76 | 0.03 |
|  | API | 12.24 | 20.94 | <0.01 | 19.35 | 13.12 | <0.01 |
|  | Hispanic | 62.44 | 45.14 | <0.01 | 46.35 | 62.84 | <0.01 |
|  | Black/African American | 16.57 | 23.99 | <0.01 | 23.79 | 16.05 | <0.01 |
|  | Multiple race | 0.36 | 0.57 | 0.48 | 0.52 | 0.4 | 0.68 |
|  | Eligible for Free Lunch | 65.87 | 55.2 | <0.01 | 54.89 | 67.28 | <0.01 |
|  | English Learner | 50.82 | 34.79 | <0.01 | 35.39 | 51.77 | <0.01 |

### Figure S11: Cumulative incidence of influenza-related hospitalizations in the intervention and comparison districts, by age and race

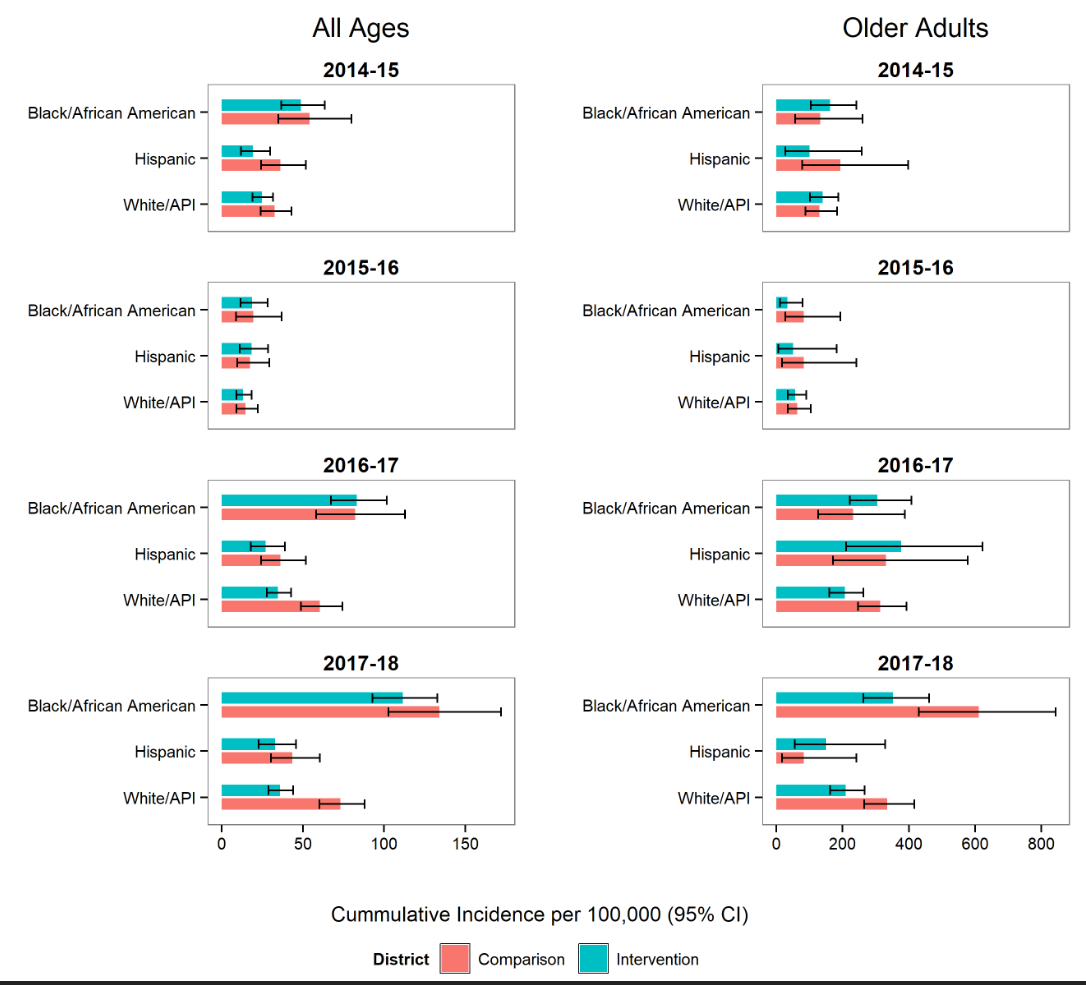

### Figure S12: Difference-in-differences in the cumulative incidence of influenza-related hospitalizations in the intervention district versus comparison district, non-elementary school ages

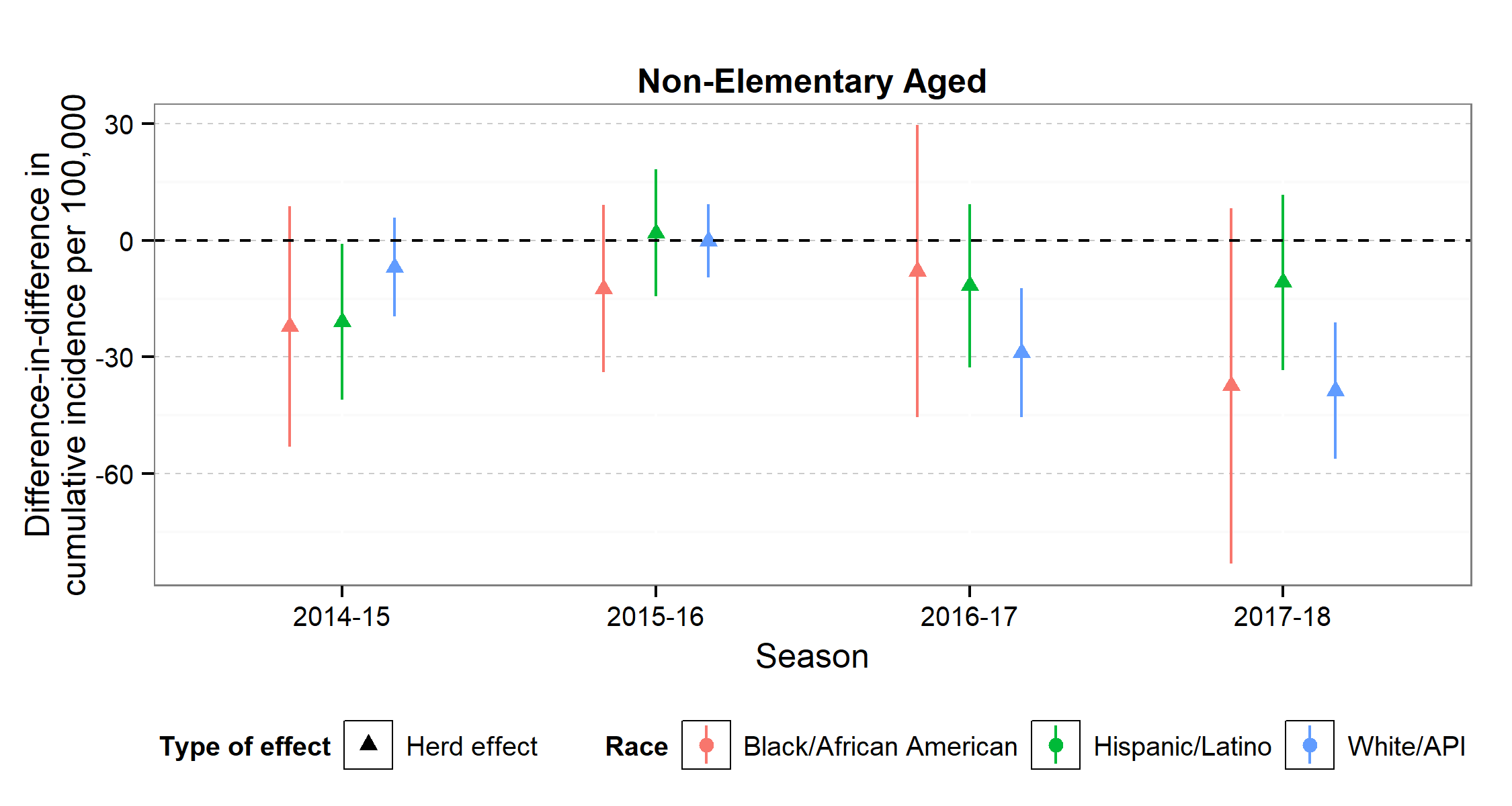
